# Epidemiology and antibiotic resistance profile of *Helicobacter pylori* infection in Cameroon: a systematic review with meta-analysis

**DOI:** 10.1101/2021.01.13.21249766

**Authors:** Corinne Raïssa Ngnameko, Jean Jacques Noubiap, Freddy Brice Simo Nemg, Mohamadou Abdou Galdima, Guy Roussel Takuissu-Nguemto, Sophie Ngo-Boumso, Frederic Nico Njayou, Paul F. Moundipa, Jean Joel Bigna

**Author notes:** **Correspondence to:** Dr. Jean Joel Bigna. Department of Epidemiology and Public Health, Centre Pasteur of Cameroon, Yaoundé, Cameroon.

## Abstract

**Objectives:** Although global epidemiology of *Helicobacter pylori* (HP) infection is well characterized, country-specific figure is more accurate for context-specific tailored interventions. The aim was to determine the prevalence, factors associated with infection, antibiotic resistance profile, and genotypes of HP in Cameroon.

**Design:** A systematic review with meta-analysis.

**Participants:** People living in Cameroon regardless of their clinical profile.

**Data sources and synthesis:** Observational studies published in PubMed, EMBASE, African Index Medicus, African Journals Online, and Health Sciences and Diseases up to October 12^th^, 2020. Study selection, data extraction, and methodological quality assessment were done by two independent authors. Random-effect meta-analysis served to pool prevalence data.

**Results:** Fifteen studies were included. None investigated the genotypes of HP. Among symptomatic patients, the most common used test, urea breath test on gastric biopsy, yielded a prevalence 57.8% (95% confidence interval (CI): 34.3-79.5) of HP infection. This prevalence significantly varied between 47.0% (95%CI: 43.0-51.0) for antigen test in stool samples and 71.2% (95%CI: 59.1-81.9) for Giemsa on gastric biopsy; *p* = 0.0006. One study in asymptomatic children reported the prevalence of 52.3% (95%CI: 44.9-59.6) using antigen test for stool samples. Among the six studies investigating factors associated with HP infection, one performed a multivariable model and identified being student as protective factor compared to being employed (Odds ratio: 0.09; 95%CI: 0.02–0.49). For antibiotics used for first line treatment, HP was resistant to amoxicillin (85.6-97.1%), metronidazole (93.2-97.9%), and clarithromycin (13.6-44.7%) as reported in two studies.

**Limitation:** Data were from six of the ten regions of Cameroon, hindering the generalizability of the findings to the country.

**Conclusion:** This study depicted a high prevalence of HP infection and a worrying resistance profile to first-line antibiotics. When waiting for well-conducted studies, updated guidelines are needed for clinical practice for first-line antibiotic treatment.

**Registration:** PROSPERO, CRD42020220084.

**Strengths and limitations of this study:**

- Not all the 10 regions of the country were represented in this study, hindering the generalizability of the findings.
- We were not able to substantially investigate sources of heterogeneity in the prevalence of *Helicobacter pylori* infection due to scarcity of data.
- Evidence investigating factors associated with *Helicobacter pylori* infection are weak and there was scarcity on data exploring antibiotic resistance profile of *Helicobacter pylori*.
- This study is the first to provide an estimate of the epidemiology and resistance profile of *Helicobacter pylori* in Cameroon.

## Introduction

*Helicobacter pylori* (*H. pylori*) is a Gram negative bacterium belonging in the family of *Helicobacteriaceae*. This bacterium has been isolated from feces, saliva and dental plaques, suggesting that transmission is possible by ingestion of food or water contaminated by the feces through the gastro-oral route or by person-to-person transmission.^1 2^ *H. pylori* infection is global public health concern with 4.4. billion people living with *H. pylori* worldwide.^3^ The global prevalence of *H. pylori* infection is 44.3%, with the prevalence higher in developing countries (50.8%) and in Africa (70.1%) compared to other regions.^3^ Although *H. pylori* infection would have a protective effect against esophageal adenocarcinoma and Barrett’s esophagus;^4^ *H. pylori* infection increases the risk for gastroesophageal reflux, peptic or duodenal ulcer, and colorectal and gastric cancers.^5-9^ In addition, *H. pylori* infection is also associated with non-gastrointestinal disorders like diabetes, obesity, acute coronary syndrome, periodontitis, nonalcoholic fatty liver, Sjögren syndrome, systemic sclerosis, pre-eclampsia, and infertility.^10-20^ Therefore, curbing the burden of these diseases requires targeted interventions to eradicate *H. pylori* infection.^9 21-23^ Antibiotic resistance profile would make difficult the eradication of *H. pylori* infection. Globally, primary and secondary resistance rates to clarithromycin, metronidazole, and levofloxacin were ≥ 15% in all WHO regions, except primary clarithromycin resistance in the Americas (10%) and South-East Asia region (10%) and primary levofloxacin resistance in the European region (11%).^23^

Although the global and regional epidemiology and antibiotic resistance profile of *H. pylori* infection is characterized, country-specific figure is more accurate for context-specific tailored interventions. To date, there is no systematic review summarizing data on the epidemiology and antibiotic resistance profile of *H. pylori* infection in Cameroon. Therefore, the aim was to determine the prevalence, factors associated with infection, antibiotic resistance profile, and genotypes of *H. pylori* infection in populations living in Cameroon. Such data would help policy makers for national and context-specific interventions, would help for advising local clinical guidelines, and highlight gap between current evidence and new research needed in the country.

## Methods

### Design

The Joanna Briggs Institute guidelines was used for the methodology of this systematic review and meta-analysis.^24^ This systematic review and meta-analysis was registered with PROSPERO, number CRD42020220084. This review was reported as per PRISMA guidelines.^25^

### Search strategy

We performed a search of PubMed, Excerpta Medica Database (EMBASE), African Index Medicus, African Journals Online, and the Cameroonian local journal Health Sciences and Diseases to identify relevant studies published on *H. pylori* in Cameroon up to October 12^th^, 2020; regardless of the language of publication. Both text words and medical subject heading terms were used. Search strategy conducted in EMBASE is shown in the Appendix (Supplementary Table 1). A manual search that consists in scanning the reference lists of eligible papers and other relevant review articles was applied.

**Table 1.** Prevalence of *Helicobacter pylori* in patients with gastro-intestinal symptoms or disease* and in children general population.

|  | Prevalence (95%CI) | N studies | N participants | I<sup>2</sup>, % | P heterogeneity | P Egger | P difference |
| --- | --- | --- | --- | --- | --- | --- | --- |
| **Symptomatic** |  |  |  |  |  |  |  |
| • Antigen test | 47.0 (43.0-51.0) | 2 | 600 | 0.0 | 0.660 | NA | 0.0006 |
| • PCR, DNA | 54.5 (47.2-61.7) | 1 | 178 | NA | NA | NA |  |
| • Urea breath test | 57.8 (34.3-79.5) | 8 | 1,298 | 98.5 | < 0.0001 | 0.047 |  |
| • Antibody test | 62.1 (46.2-76.8) | 4 | 1,190 | 96.4 | < 0.0001 | 0.074 |  |
| • Giemsa | 71.2 (59.1-81.9) | 1 | 59 | NA | NA | NA |  |
| **Asymptomatic children** |  |  |  |  |  |  |  |
| • Antigen test | 52.3 (44.9-59.6) | 1 | 176 | NA | NA | NA |  |

PCR: polymerase chain reaction, DNA: deoxyribonucleic acid, NA: not applicable.
\* include dyspepsia, gastritis, gastric pain, gastroduodenal ulcer,

### Eligibility criteria

We considered observational (cross-sectional, case-control, and cohort) studies conducted in general population and populations with specific conditions living in Cameroon. *H. pylori* infection had to be laboratory-diagnosed. Studies had to report the prevalence (or enough data to compute this estimate) or factors associated with *H. pylori* infection, or had to investigate genotypes or the relevant antibiotic (azithromycin, clarithromycin, erythromycin, metronidazole, tetracycline, minocycline, ampicillin, amoxicillin (+ clavulanic acid), levofloxacin, ciprofloxacin, levofloxacin) resistance profile of *H. pylori*.^23^ We have not considered studies in which patients self-reported the infection. Studies conducted in Cameroonian living outside Cameroon were not considered.

### Study selection

Two review authors independently screened records based on titles and abstracts for eligibility. Full texts of articles deemed potentially eligible were retrieved. Further, these review authors independently assessed the full text of each study for final inclusion. Disagreements when existing were solved through discussion and consensus.

### Data extraction and management

Data were extracted using a preconceived data abstraction form. Two review authors independently extracted data including: name of the first author, publication year, study design, setting, sampling method, samples collection period, timing of data collection and analysis, site of recruitment, underlying conditions, number of patients screened, number of patients infected with *H. pylori*, diagnostic techniques used, laboratory samples, genotype profile, antibiotic resistance profile, mean age, factors associated with *H. pylori* infection, and proportion of male participants. Disagreements between review authors were reconciled through consensus.

Two review authors assessed the risk of bias in included studies using the tool developed by the Joanna Briggs Institute. Disagreements were solved through discussion and consensus. The following items were considered: sampling method (probabilistic sampling *vs*. non-probabilistic sampling), diagnostic method of *H. pylori* (direct test [stool antigen test, urea breath test of gastric biopsy samples, culture of gastric biopsy samples, stool culture, PCR] vs. indirect test [antibody detection, saliva, antigen detection]),^26^ precision (acceptable vs. low), same procedure for diagnostic of *H. pylori* (yes vs. no).

### Data synthesis and analysis

Data analyses used the *‘meta’* packages of the statistical software R (version 3.6.3, The R Foundation for statistical computing, Vienna, Austria).^27^ Single arcsine transformation with random-effects meta-analysis model was used to pool prevalence data of *H. pylori* infection.^28^ Egger’s test served to assess the presence of publication bias.^29^ A *p* < 0.10 on Egger test was considered indicative of statistically significant publication bias.

Heterogeneity was evaluated by the χ^2^ test on Cochran’s Q statistic,^30^ which was quantified *I*^2^ values.^31^ The *I*^2^ statistic estimates the percentage of total variation across studies due to true between-study differences rather than chance. In general, *I*^2^ values greater than 60-70% indicate the presence of substantial heterogeneity. We performed a narrative synthesis for antibiotic resistance and factors associated with *H. pylori* infection because of the heterogeneity in reporting theses outcomes and scarcity of data.

## Results

### Study selection and characteristics

We initially identified 77 records and finally retained 15 studies (Figure 1).^32-46^ Agreement between investigators for report selection based on title and abstract was κ = 0.88, and κ = 1 for final inclusion. Individual characteristics of included studies are in the Supplementary Table 2.

**Table 2.**
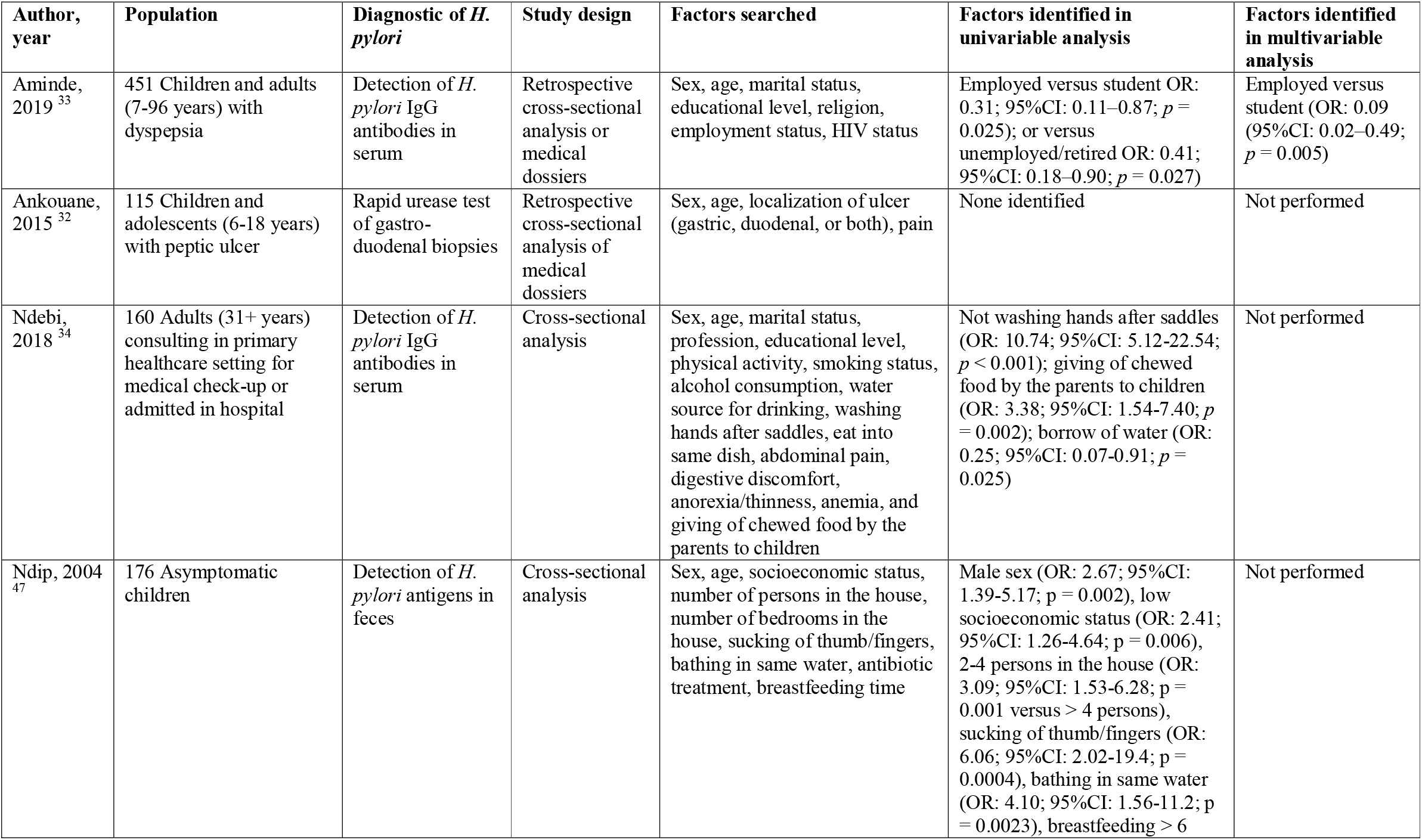
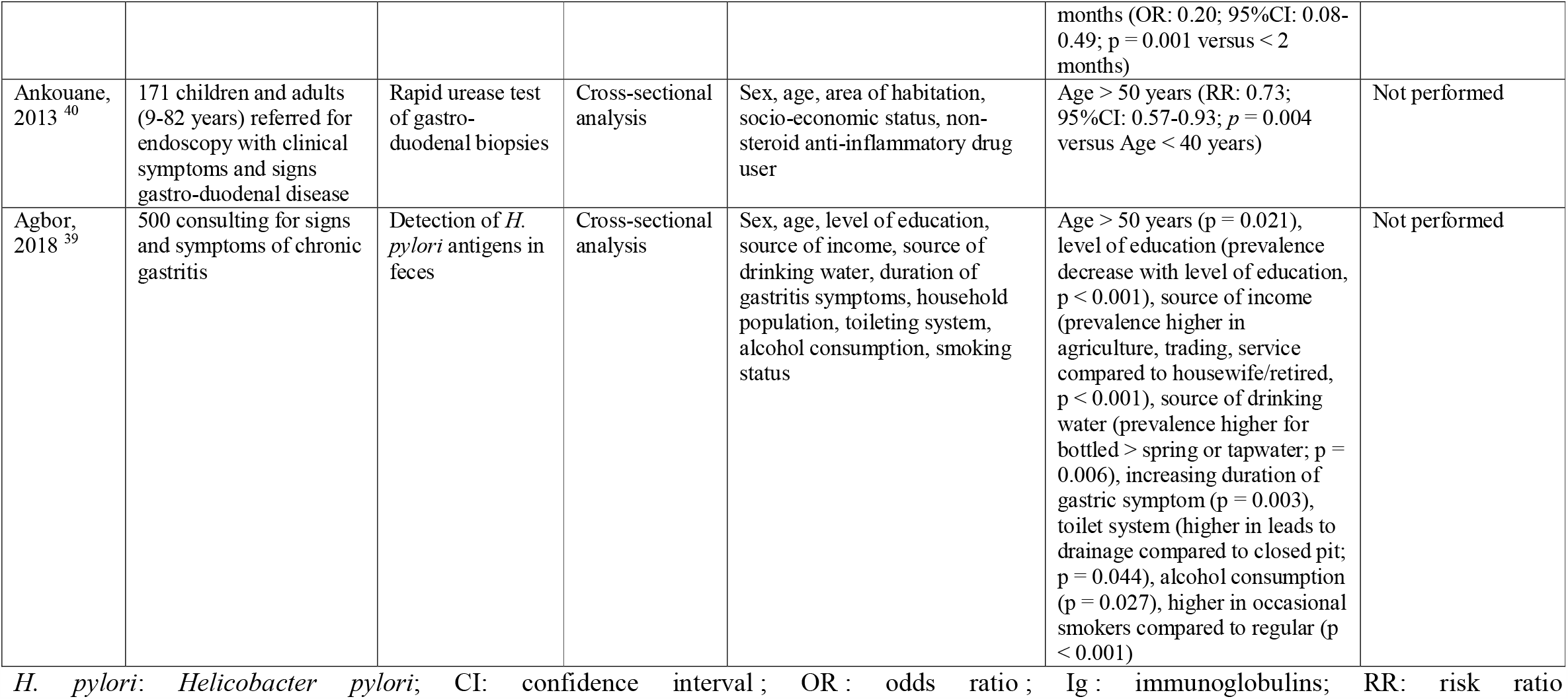
Factors associated with Helicobacter pylori infection in populations living in Cameroon.

**Figure 1.**
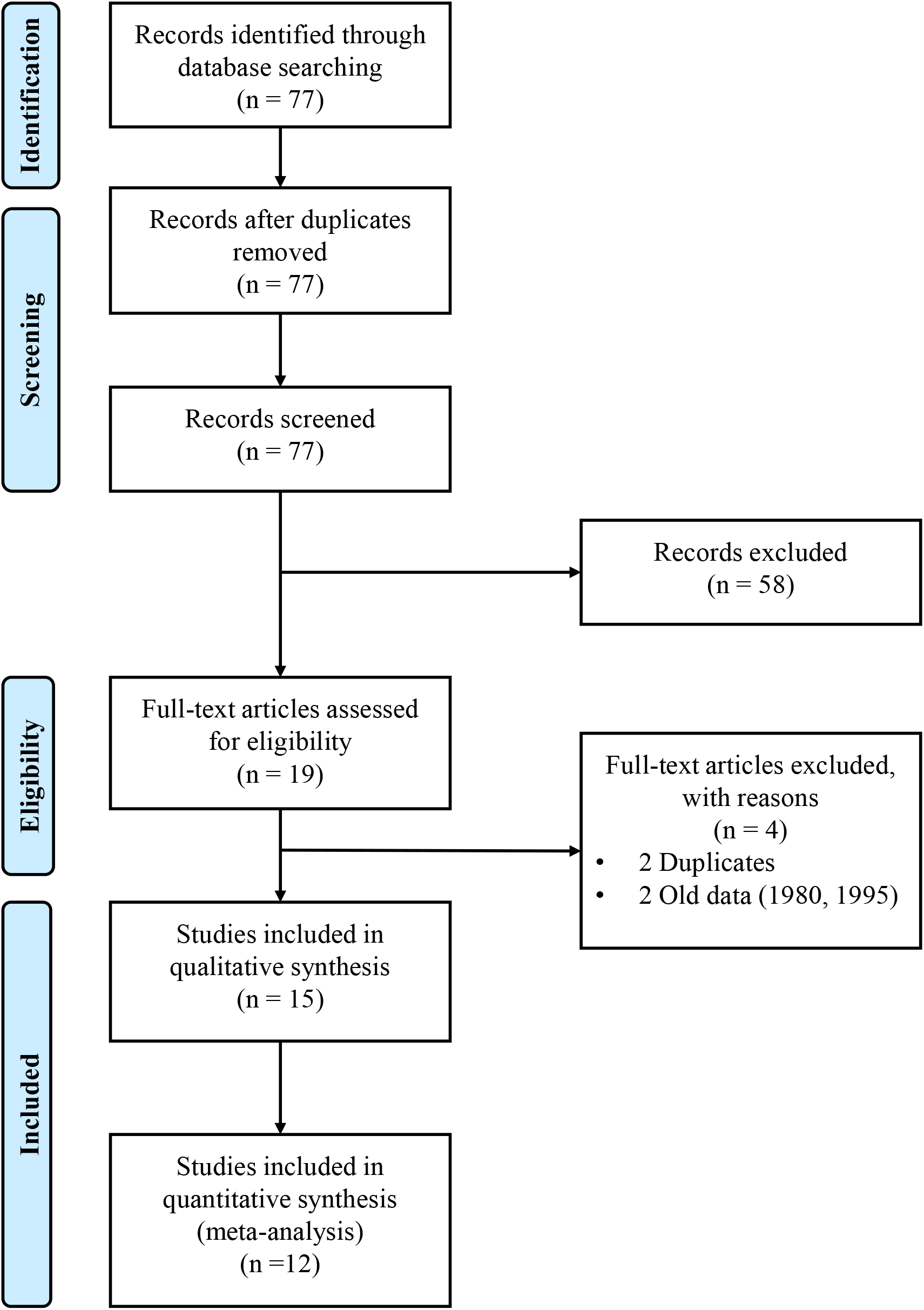
Study flow diagram.

**Figure 2.**
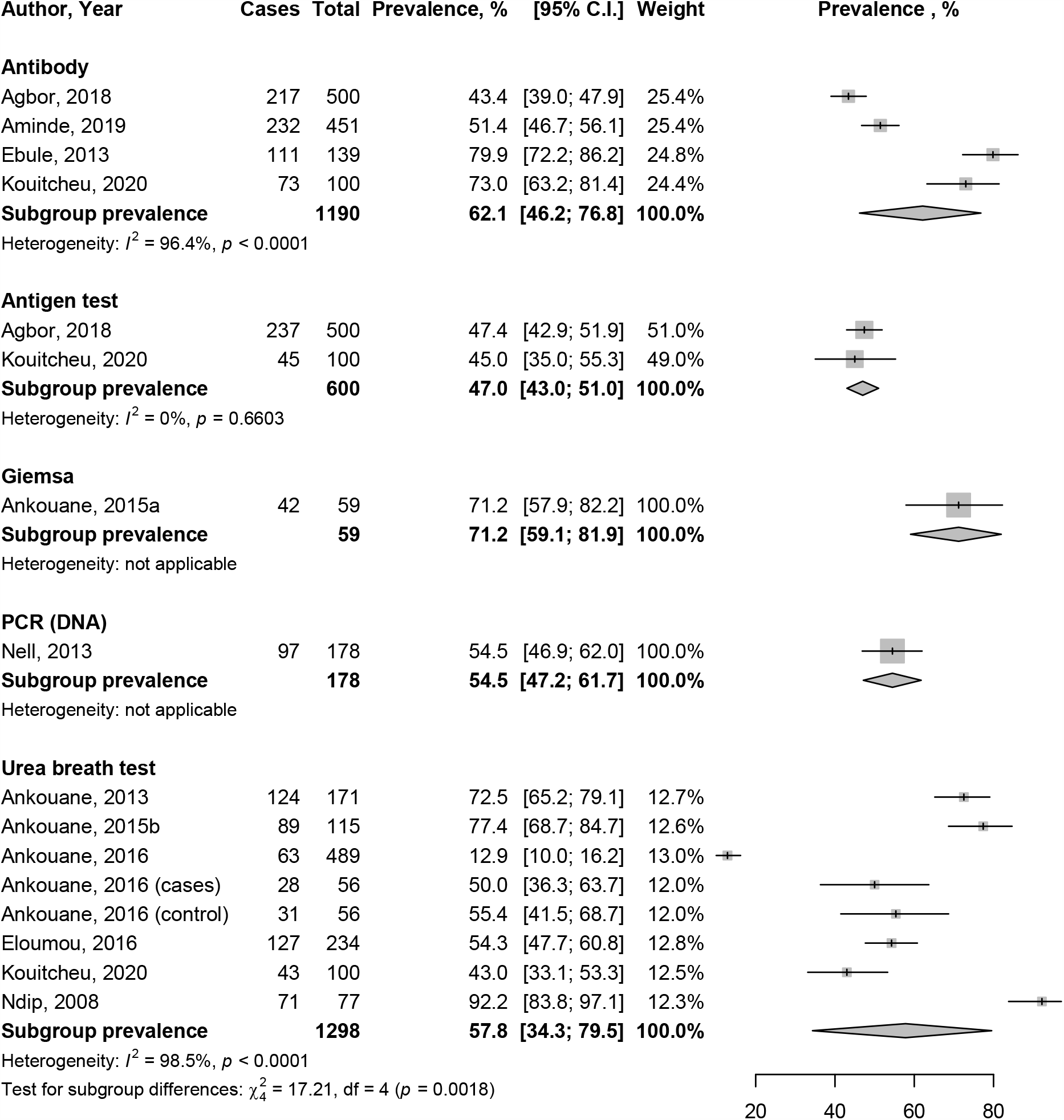
Prevalence of *Helicobacter pylori* in Cameroon according to different diagnostic methods in symptomatic patients. PCR: polymerase chain reaction, DNA: deoxyribonucleic acid

Studies were published between 2013 and 2019 and data were collected between 2006 and 2018. Fourteen studies were cross-sectional, and one was case control. Fourteen studies were hospital-based, and one was community-based. Data were from six of the ten regions of the country: Centre, Littoral, East, North-West, South-West, and West. Gastric biopsy was done in 10 studies, blood collection in five studies, and stool collection in three studies. Urea breath test were used in eight studies, antibodies were searched in five studies, antigens in three studies, Giemsa in two studies, and PCR in one study. There were fourteen studies reporting prevalence data, four studies that investigated factors associated with *H. pylori* infection, and two that investigated antimicrobial resistance. None of the studies investigated the genotypes of *H. pylori*. One study was conducted among asymptomatic general population of children and the remaining were conducted among symptomatic patients.

Two studies performed probabilistic sampling and the remaining non-probabilistic sampling. Twelve studies prospectively analysed data and the remaining retrospectively. The response rate was acceptable in 12 studies and not reported in three studies. (Supplementary Table 2).

### Prevalence of *Helicobacter pylori* in Cameroon

Twelve studies investigated the prevalence of *H. pylori* infection. Among symptomatic people,^32 33 35 37-45^ the most common used test, urea breath test on gastric biopsy, yielded a prevalence 57.8% (95%CI: 34.3-79.5), with substantial heterogeneity (Table 1). The prevalence of *H. pylori* infection significantly varied between 47.0% (95%CI: 43.0-51.0) for antigen test in stool samples and 71.2% (95%CI: 59.1-81.9) for Giemsa on gastric biopsy (Figure 1), with significant difference between diagnostic methods, *p* = 0.0006 (Table 1). The publication bias was marginal (Table 1).

One study reported the prevalence of *H. pylori* in general population.^46^ The study was conducted among 176 asymptomatic children on stool samples. The prevalence was 52.3% (95%CI: 44.9-59.6) using antigen test.

### Factors associated with *Helicobacter pylori* infection in Cameroon

Six studies investigated factors associated with *H. pylori* in symptomatic patients and in general population of children and adults (Table 2).^32-34 39 40 46^ Aminde and colleagues in a retrospective cross-analysis of 451 children and adults with dyspepsia, identified being student (odds ratio [OR]: 0.41; 95%CI: 0.18-0.90) and unemployed/retired (OR: 0.31; 95%CI: 0.11-0.87) as protective factor compared to employed, as well as being HIV-infected (OR: 0.49; 95%CI: 0.15-0.68). ^33^ In the multivariable model, only being student remained associated with *H. pylori* immunoglobulins G seropositivity (Table 2).^33^

The remaining studies did not perform multivariable analysis. Ankouane and colleagues in a 9-year retrospective analysis of 115 children and adolescents with peptic ulcer disease did not identify any factor associated with *H. pylori* infection on gastroduodenal biopsies using rapid urease test.^32^ Ndebi and colleagues in a cross-sectional study of 160 adults consulting in a primary healthcare setting for medical check-up or with hospital admission identified not washing hands after saddles (OR: 10.74; 95%CI: 5.12-22.54), giving of chewed food by the parents to children (OR: 3.38; 95%CI: 1.54-7.40), and borrow water (OR: 0.25; 95%CI: 0.07-0.91).^34^ Ndip and colleagues in a cross-sectional study of 176 asymptomatic children identified male sex (OR: 2.67; 95%CI: 1.39-5.17), low socioeconomic status (OR: 2.41; 95%CI: 1.26-4.64), 2-4 persons in the house (OR: 3.09; 95%CI: 1.53-6.28 versus > 4 persons), sucking of thumb/fingers (OR: 6.06; 95%CI: 2.02-19.4), bathing in shared water (OR: 4.10; 95%CI: 1.56-11.2), breastfeeding > 6 months (OR: 0.20; 95%CI: 0.08-0.49; versus < 2 months).^46 47^ Ankouane and colleagues in a cross-sectional study of 171 children and adults identified age > 50 years (RR: 0.73; 95%CI: 0.57-0.93) as factor associated with *H. pylori* infection with rapid urease test on gastric biopsies.^40^ Agbor and colleagues in a cross-sectional study of feces from 500 patients consulting for signs and symptoms of chronic gastritis identified age > 50 years (p = 0.021), level of education (prevalence decrease with level of education, p < 0.001), source of income (prevalence higher in agriculture, trading, service compared to housewife/retired, p < 0.001), source of drinking water (prevalence higher for bottled > spring or tap water; p = 0.006), increasing duration of gastric symptom (p = 0.003), toilet system (higher in leads to drainage compared to closed pit; p = 0.044), alcohol consumption (p = 0.027), smoking status (higher in occasional smokers compared to regular; p < 0.001) associated detection of *H. pylori* antigen.^39^

### Antibiotic resistance profile of *Helicobacter pylori* in Cameroon

Two studies reported antibiotic resistance profile of *H. pylori* (Table 3).^35 36^ Ndip and colleagues analysed 132 gastric biopsy specimens for *H. pylori* resistance against four antibiotics (clarithromycin, tetracycline, amoxicillin, and metronidazole). The proportion of resistance varied from 43.9% for tetracycline to 93.2% for metronidazole.^35^ In a study by Kouitcheu Mabeku and colleagues in 140 gastric biopsy specimens,^36^ the authors did not find any resistance against azithromycin, ciprofloxacin, and levofloxacin. The resistance was less than 20% for three antibiotics (minocycline, tetracycline, and clarithromycin). The resistance was 47.9% for erythromycin. For the remaining antibiotics, the resistance was higher than 95% (amoxicillin, metronidazole, ampicillin (+ clavulanic acid)).

**Table 3.** Antibiotic resistance profile of *Helicobacter pylori* in Cameroon.

| Author | Sample | Description of antibiotics resistance |
| --- | --- | --- |
| Kouitcheu Mabeku, 2019 <sup>36</sup> | 140 gastric biopsy specimens | Azithromycin 0/140; 0%<br>Ciprofloxacin 0/140; 0%<br>Levofloxacin 0/140; 0%<br>Minocycline 1/140; 0.7%<br>Tetracycline 4/140; 2.9%<br>Clarithromycin 19/140; 13.6%<br>Erythromycin 67/140; 47.9%<br>Amoxicillin 136/140; 97.1%<br>Metronidazole 137/140; 97.9%<br>Ampicillin 140/140; 100%<br>Amoxicillin + clavulanic acid 140/140; 100% |
| Ndip, 2008 <sup>35</sup> | 132 gastric biopsy specimens | Tetracycline 58/132; 43.9%<br>Clarithromycin 59/132; 44.7%<br>Amoxicillin 113/132; 85.6%<br>Metronidazole 123/132; 93.2% |

## Discussion

This systematic review with meta-analysis of 15 studies depicted a high prevalence of *H. pylori* infection among different people living in Cameroon using different diagnostic methods. There was a high variability among reported factors associated with this infection. *H. pylori* was resistant to most of antibiotics at 50% level or more.

The prevalence of *H. pylori* infection we found in this study is in the range of estimates reported from other countries of Africa (46.8-87.7%) as well as for other regions of the world (44.3%, 95% CI: 40.9-47.7).^3 48^ At the turn of the 21^st^ century, the prevalence of *H. pylori* has been declining in highly industrialized countries of the Western world, whereas prevalence has plateaued at a high level in developing (like in Cameroon) and newly industrialized countries.^3^ The high prevalence in the country may be explained by poor access to adequate sanitation, poor access to clean water, and low socioeconomic status known to be associated with *H. pylori* infection. The overall situation is concerning in the country. For instance, few (51%) family had access to enough water, a fifth of the communities used exclusively unimproved water sources and 52% mixes both unimproved and improved water sources, 31% of the community do not treat drinking water, and only few (67%) people have soap at home and even less have handwashing facilities (63%).^49^ Therefore, curbing the burden of *H. pylori* infection required a broad intervention for improving hygiene, sanitation, and water supply profile in the country. Performing systematic and community testing for *H. pylori* could be explored, however, in a resource-limited setting with high burden of other diseases seems to be unrealistic. A focus should be done for patient with high risk of infection; for instance, those with gastroduodenal symptoms.

At individual level, several factors have been investigated with *H. pylori* infection; however, only one study performed a multivariable model identifying being student as a protective factor compared to being employed. Multiple factors have been identified in the univariable model, however it is difficult to exploit these findings and make recommendations. Therefore, high-quality and well-conducted community-based epidemiological studies are needed to identify risk factors associated with *H. pylori* infection in the country. Such studies would help to identify country-specific risk factors of *H. pylori* infection and adopt related and tailored recommendations.

In Cameroon, first line treatment for *H. pylori* include two main strategies: concomitant quadruple therapy (proton pump inhibitor + amoxicillin + metronidazole + clarithromycin for 14 days) and sequential quadruple therapy (proton pump inhibitor + amoxicillin for 5 days, followed by proton pump inhibitor + metronidazole + clarithromycin for 5 days).^50^ As reported in a study conducted among 165 patients, the most used is the first one (63%).^50^ From this study, the treatment success rate of these strategies were 73.9% without significant difference between them.^50^ However, using these antibiotics in this context is questionable looking at the findings of the two studies on antibiotic resistance we found. *H. pylori* was resistant to amoxicillin (85.6-97.1%), metronidazole (93.2-97.9%), and clarithromycin (13.6-44.7%). Only clarithromycin had profile resistance < 50%. Therefore, all treatment initiation against *H. pylori* should be guided by the antibiotic resistance profile. However, in a resource-limited setting where the resistance profile test is only (but not all) accessible in secondary/tertiary healthcare facilities, this strategy is unrealistic. Therefore, two strategies should be adopted, one guided by antibiotic resistance profile and the second one based on probabilistic nonresistance profile. The last one requires a broad analysis of *H. pylori* antibiotic resistance profile considering actual evidence and performing a nationally-representative and community-based study to design tailored recommendations for context-specific probabilistic and efficacious first-line antibiotic treatment.

The findings of this study should be interpreted with caution. Not all the 10 regions of the country were represented in this study hindering the generalizability of the findings of this study to the entire country. We were not able to substantially investigate sources of heterogeneity in the prevalence of *H. pylori* infection due to scarcity of data, although heterogeneity is practically inevitable in a meta-analysis of prevalence studies. In the absence of a nationally representative population-based study taking into account all regions of the country, this study is the first to provide an estimate of the epidemiology and resistance profile of *H. pylori* in Cameroon without a lot of financial, logistical, and material expenses in the context of a resource-limited country.

This study depicted a high prevalence of *H. pylori* infection in both symptomatic and asymptomatic populations. This study also highlights a worrying resistance profile to first-line antibiotics. When waiting for well-conducted studies to identify risk factors associated with *H. pylori* infection and to have a more accurate profile of antibiotic resistance profile, updated guidelines are needed for clinical practice for first-line treatment in the country.

## Data Availability

All data relevant to the study are included in the article or uploaded as supplementary information.

## Author contributions

Conception and design: JJB, MAG, CRN, FNN, FBNS. Search strategy: JJB. Study selection: JJB, FBNS. Data extraction: JJB, CRN, FBNS. Data analysis: JJB and JJN. Manuscript drafting: JJB, CRN, JJN. Manuscript revision: JJB, MAG, PFM, CRN, SNB, FNN, JJN, FBNS, GRTN. Guarantor of the review: JJB. Approved the final version of the manuscript: All authors.

## Acknowledgements

None.

