## Appendix for "Epidemiology and antibiotic resistance profile of *Helicobacter pylori* infection in Cameroon: a systematic review with meta-analysis"

---

**Supplementary Table 1. Search strategy in EMBASE**

| Search | Search terms |
|---|---|
| #1 | 'helicobacter pylori'/exp OR 'helicobacter pylori' OR 'helicobacter'/exp OR helicobacter |
| #2 | 'cameroon'/exp OR cameroon OR 'cameroun'/exp OR cameroun |
| #3 | #1 AND #2 |

**Supplementary Table 2. Characteristics of included studies**

| Author | Year of publication | Design | Period | Sampling | Timing | Response | Setting | Cities | Samples | Laboratory test | %Males | Age, years | Population characteristics | Sample |
|---|---|---|---|---|---|---|---|---|---|---|---|---|---|---|
| Ankouane | 2013 | Cross-sectional | 2012-2013 | Convenience | Prospectively | > 80% | Hospital | Yaounde | Gastric Biopsy | Urea breath test | 41 | 42.3 | Gastro-duodenal pathologies | 171 |
| Ankouane | 2015 | Case control | 2014-2015 | Consecutive | Prospectively | NR | Hospital | Yaoundé and Douala | Gastric Biopsy | Urea breath test | 48 | 42.8 | Gastro-Intestinal symptoms, HIV+ and HIV- | 112 |
| Nell | 2015 | Cross-sectional | NR | Convenience | Prospectively | > 80% | Hospital | Abong Mbang, Yokadouma | Gastric Biopsy | PCR (DNA) | 33 | | Gastric Pain | 178 |
| Ndip | 2015 | Cross-sectional | 2006 | Convenience | Prospectively | > 80% | Hospital | Douala | Gastric Biopsy | Urea breath test | 45 | 44.5 | Gastro-duodenal pathologies | 77 |
| Ndip | 2015 | Cross-sectional | NR | Random | Prospectively | > 80% | Community | Buea, Limbe | Stool | Antigen test | 49 | 4.3 | Asymptomatic Children | 176 |
| Ankouane | 2015 | Cross-sectional | 2013-2014 | Convenience | Prospectively | > 80% | Hospital | Younde | Gastric Biopsy | Giemsa | 43 | 43 | Chronic antral gastritis | 59 |
| Ebule | 2015 | Cross-sectional | 2008-2009 | Convenience | Prospectively | > 80% | Hospital | Tombel | Blood | Antibody | 33 | 46.7 | Dyspepsia | 139 |
| Eloumou | 2015 | Cross-sectional | 2013 | Convenience | Prospectively | > 80% | Hospital | Douala | Gastric Biopsy | Urea breath test | 42 | 43.7 | Gastro-duodenal lesions | 234 |
| Kouitcheu | 2020 | Cross-sectional | 2017-2018 | Consecutive | Prospectively | > 80% | Hospital | Douala | Blood, Stool, Gastric biopsy | Antibody, Antigen, Urea breath test | 34 | 41.5 | Dyspepsia | 100 |
| Kouitcheu | 2015 | Cross-sectional | 2013-2015 | Consecutive | Prospectively | > 80% | Hospital | Douala | Gastric Biopsy | Gram staining, Urea breath test, catalase/oxidase tests | NR | NR | NR | 140 |
| Ndebi | 2015 | Cross-sectional | 2016 | Convenience | Prospectively | > 80% | Hospital | Dschang | Blood | Antibody | NR | NR | General consulting primary care setting | 160 |
| Ankouane | 2015 | Cross-sectional | 2006-2014 | Systematic | Retrospectively | > 80% | Hospital | Yaounde | Gastric Biopsy | Urea breath test | 73 | 14.7 | Peptic ulcer Children | 115 |
| Ankouane | 2016 | Cross-sectional | 2011-2015 | Convenience | Retrospectively | NR | Hospital | Yaoundé | Gastric Biopsy | Urea breath test | 64 | 48.9 | Gastro-duodenal Ulcer | 489 |
| Agbor | 2018 | Cross-sectional | 2013-2015 | Convenience | Prospectively | NR | Hospital | Melong | Stool, Blood | Antigen test, Antibody | 67 | | Gastritis | 500 |
| Aminde | 2019 | Cross-sectional | 2012-2016 | Time-Location | Retrospectively | > 80% | Hospital | Wum | Blood | Antibody | 36 | 40.7 | Dyspepsia | 451 |

NR: Not reported.
